# Initial vancomycin versus metronidazole for the treatment of first-episode non-severe *Clostridioides difficile* infection

**DOI:** 10.1101/2020.12.04.20243766

**Authors:** Kevin Zhang, Patricia Beckett, Salaheddin Abouanaser, Marek Smieja

**Affiliations:** Faculty of Medicine, University of Toronto, Toronto, Ontario, M5S 1A8, Canada; St. Joseph’s Healthcare Hamilton, Hamilton, Ontario, L8N 4A6, Canada; Department of Pathology and Molecular Medicine, McMaster University, Hamilton, Ontario, L8S 4L8, Canada

**Keywords:** *Clostridioides difficile*, *Clostridium difficile*, vancomycin, metronidazole, recurrence, incidence, mortality

## Abstract

**Objective:** *Clostridioides difficile* infection (CDI) is the leading cause of infectious nosocomial diarrhea. Although initial fidaxomicin or vancomycin treatment is recommended by most major guidelines to treat severe CDI, there exists varied recommendations for first-episode non-severe CDI. Given the discrepancy in current treatment guidelines, we sought to evaluate the use of initial vancomycin versus metronidazole for first-episode non-severe CDI.

**Methods:** We conducted a retrospective cohort study of all adult inpatients with first-episode CDI at our institution from January 2013 to May 2018. The initial vancomycin versus initial metronidazole cohorts were examined using a multivariate logistic regression model.

**Results:** Patients (n = 737) had a median age of 72.3 years and 357 (48.4%) had hospital-acquired infection. Among patients with non-severe CDI (n = 326), recurrence, new incident infection, and 30-day mortality rates were 16.2%, 10.9%, and 5.3%, respectively, when treated with initial metronidazole, compared to 20.0%, 1.4%, and 10.0%, respectively, when treated with initial vancomycin. In an adjusted multivariable analysis, the use of initial vancomycin for the treatment of non-severe CDI was associated with a reduction in new incident infection (OR_adj_: 0.11; 95% CI: 0.02–0.86; P=0.035), compared to initial metronidazole.

**Conclusions:** Initial vancomycin was associated with a reduced rate of new incident infection in the treatment of adult inpatients with first-episode non-severe CDI. These findings support the use of initial vancomycin for all inpatients with CDI, when fidaxomicin is unavailable.

## Introduction

*Clostridioides difficile* infection (CDI) manifests itself in patients with an altered gut microbiota, and is commonly caused by exposure to antibiotics.^1^ As the leading cause of infectious nosocomial diarrhea, CDI continues to be a major burden within healthcare institutions and in the community.^1^ Despite undergoing lengthy courses of antimicrobial therapy, however, recurrence is common, with approximately 20% of patients experiencing debilitating episodes of repeated disease.^2^ Recurrent CDI is associated with a greater risk for future recurrence and severe complications such as hypotension, perforation, and toxic megacolon.^2^ Therefore, early interventions aimed at preventing recurrent CDI and reinfection are needed to improve outcomes, decrease the number of patient days spent in isolation, and improve quality of life.

St. Joseph’s Healthcare Hamilton is a multi-site teaching hospital located in Hamilton, Ontario, Canada with a dedicated *Clostridioides difficile* consultation service for inpatients and referred outpatients. While the consultation team may offer fecal microbiota transplantation to patients with multiple CDI episodes or refractory disease,^3,4^ clinical interventions for first-episode CDI primarily involve the use of antibiotics. In the past, metronidazole was indicated for non-severe CDI at our institution, while vancomycin was reserved for febrile patients with elevated white counts or for recurrent episodes. In recent years, however, there has been a shift to initial vancomycin to treat first-episode non-severe CDI at our institution, which is consistent with guidelines published by the Infectious Diseases Society of America (IDSA) and Society for Healthcare Epidemiology of America (SHEA), when fidaxomicin is unavailable.^5^ Importantly, guidelines published by the European Society of Clinical Microbiology and Infectious Diseases, the World Society of Emergency Surgery, and the Australasian Society of Infectious Diseases still recommend the use of metronidazole for the first episode of non-severe disease.^6–8^

Although initial fidaxomicin or vancomycin treatment is recommended by most major guidelines to treat severe CDI, there exists a discrepancy in treatment recommendations for first-episode non-severe CDI.^5–8^ There have been limited studies with long-term follow-up evaluating the use of initial vancomycin compared to initial metronidazole to treat non-severe inpatient CDI. In two large randomized controlled trials with follow-up of 3 weeks and 4 weeks, respectively, subgroup analyses stratified by severity did not demonstrate significant differences in relapse rates between the vancomycin and metronidazole treatment groups for non-severe CDI.^9,10^ Similarly, two cohort studies reported that relapse rates and 30-day mortality did not differ among patients with mild-to-moderate CDI treated with metronidazole or vancomycin.^11,12^ Therefore, we sought to identify predictors of adverse clinical outcomes and evaluate the use of initial vancomycin versus metronidazole for the treatment of first-episode non-severe CDI among adult inpatients.

## Methods

### Study population

We conducted a retrospective cohort study of all adult inpatients with first-episode CDI at our institution from January 1, 2013 to May 31, 2018. Patients were identified from hospital infection control registries maintained by Infection Prevention and Control, and all entries were validated against data reported to the Ontario Ministry of Health and Long-Term Care. A CDI episode was characterized clinically as 3 or more type 5–7 diarrheal stools on the Bristol stool scale, together with a positive *Clostridioides difficile* toxin gene loop-mediated isothermal amplification assay.^13^ Patients with a prior history of CDI were excluded from the study. Similarly, colonized patients who had a positive toxin assay but did not meet the clinical criteria of 3 or more type 5–7 loose stools were excluded.

Data abstracted included: demographics, specimen collection date, date of symptom onset, source of acquisition, unit in which CDI was first diagnosed (medical, surgical, or nephrology), clinical indicators at the time of diagnosis, CDI treatment, number of isolation days, and outcomes including: recurrence, new incident infection, and death. Time to treatment and time to recurrence were measured from the start of the initial CDI diagnosis. Hospital-acquired infection (HAI) was defined as CDI present >72 hours after admission, whereas non-HAI was defined as CDI present ≤72 hours after admission.^14^ CDI severity was defined according to IDSA and SHEA guidelines.^13^ A patient with white blood cell count ≥15 × 10^9^/L or a serum creatinine level >1.5 mg/dL was considered to have severe CDI.^13^

### Exposures and outcomes

The primary exposures in this study were the treatment of first-episode non-severe CDI with initial vancomycin or metronidazole. The outcomes of interest were recurrence, new incident infection, and all-cause 30-day mortality. All patients were followed for 26 weeks from the date of their initial CDI diagnosis. Recurrence was defined as a CDI case occurring after an episode with positive assay result in the previous 2–8 weeks, while new incident infection was defined as a CDI case occurring after more than 8 weeks.^13^ All-cause 30-day mortality was defined as death for any reason within 30 days of CDI diagnosis.

### Statistical analysis

Logistic regression was used to determine prognostic factors for the composite outcome of recurrence or all-cause 30-day mortality for the overall cohort. Recurrence was selected as a patient-important outcome, as it is associated with a greater risk for complications and adverse outcomes.^2^ All-cause 30-day mortality was included in the composite outcome as it is a potential competing event for recurrence. All factors were determined a priori based on published guidelines.^6–8,13^ Univariable analysis with P<0.2 was used, followed by stepwise selection in a multivariable logistic regression model. The number of variables entered into the model enabled 80% study power with 15 events per factor.

To evaluate the impact of initial vancomycin treatment on adverse clinical outcomes in patients with first-episode non-severe CDI, the initial vancomycin versus initial metronidazole cohorts were first examined in an unadjusted logistic regression analysis for any combination of recurrence, new incident infection, and all-cause 30-day mortality. Prognostic factors for the overall cohort were then added to the adjusted multivariable analysis.

## Results

A total of 737 inpatients with first-episode CDI were included in our study. Their demographics, clinical data at the time of CDI diagnosis, initial CDI treatment, and outcomes are summarized in Table 1. Patients had a median age of 72.3 years (Q1: 61.2, Q3: 83.3) and 326 (44.2%) were classified to have non-severe CDI. 357 (48.4%) patients were classified to have hospital-acquired infection, while 309 (41.9%) and 71 (9.6%) were classified to have acquired CDI in the community and other institutions, respectively. Most patients, 496 (67.3%), were initially diagnosed with CDI in a medical setting. At the time of CDI diagnosis, patients had an average temperature of 37.1°C (SD: 0.8) and a median white blood cell count of 12.3 × 10^9^/L (Q1: 8.7, Q3: 18.1). The median time to treatment following CDI diagnosis was 2 days (Q1: 1, Q3: 5). Most patients, 539 (73.1%), received initial metronidazole treatment, while 130 (17.6%) received initial vancomycin. Patients experienced a recurrence rate of 14.5%, a new incident infection rate of 7.3%, and a 30-day mortality rate of 12.8%. The median time to recurrence was 27 days (Q1: 22, Q3: 36) and the median length of time spent in isolation was 17 days (Q1: 7, Q3: 34).

**Table 1.**
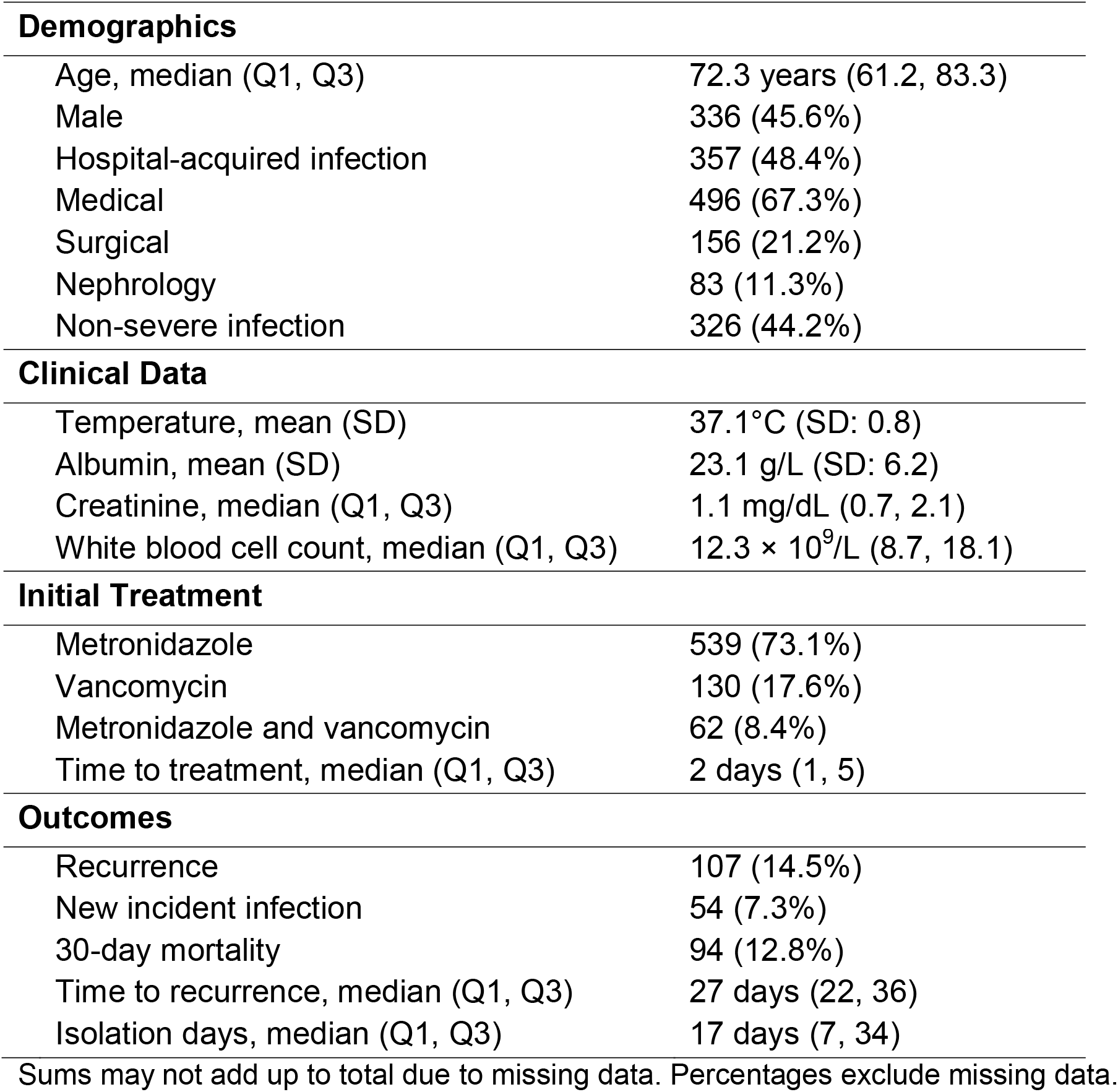
Description of patients with *Clostridioides difficile* infection (n = 737).

| **Demographics** |  |
| --- | --- |
| Age, median (Q1, Q3) | 72.3 years (61.2, 83.3) |
| Male | 336 (45.6%) |
| Hospital-acquired infection | 357 (48.4%) |
| Medical | 496 (67.3%) |
| Surgical | 156 (21.2%) |
| Nephrology | 83 (11.3%) |
| Non-severe infection | 326 (44.2%) |
| **Clinical Data** |  |
| Temperature, mean (SD) | 37.1°C (SD: 0.8) |
| Albumin, mean (SD) | 23.1 g/L (SD: 6.2) |
| Creatinine, median (Q1, Q3) | 1.1 mg/dL (0.7, 2.1) |
| White blood cell count, median (Q1, Q3) | 12.3 × 10<sup>9</sup>/L (8.7, 18.1) |
| **Initial Treatment** |  |
| Metronidazole | 539 (73.1%) |
| Vancomycin | 130 (17.6%) |
| Metronidazole and vancomycin | 62 (8.4%) |
| Time to treatment, median (Q1, Q3) | 2 days (1, 5) |
| **Outcomes** |  |
| Recurrence | 107 (14.5%) |
| New incident infection | 54 (7.3%) |
| 30-day mortality | 94 (12.8%) |
| Time to recurrence, median (Q1, Q3) | 27 days (22, 36) |
| Isolation days, median (Q1, Q3) | 17 days (7, 34) |

Sums may not add up to total due to missing data. Percentages exclude missing data.

### Overall cohort

The results of the multivariable logistic regression model to determine prognostic factors for the composite outcome of recurrence or all-cause 30-day mortality for the overall cohort are summarized in Table 2. Predictors of the composite outcome were: age ≥65 years (OR_adj_: 2.02; 95% CI: 1.37–2.96; P<0.001), hospital-acquired infection (OR_adj_: 1.98; 95% CI: 1.41–2.77; P<0.001), and white blood cell count ≥15 × 10^9^/L (OR_adj_: 1.64; 95% CI: 1.16–2.30; P=0.005).

**Table 2.** Multivariable predictors of the composite outcome of recurrence or 30-day mortality for patients in the overall cohort with *Clostridioides difficile* infection.

| **Variable** | **Odds Ratio (95% CI)** | **P** |
| --- | --- | --- |
| Age ≥ 65 years | 2.02 (1.37 – 2.96) | < 0.001 |
| Hospital-acquired infection | 1.98 (1.41 – 2.77) | < 0.001 |
| White blood cell count ≥ 15 × 10<sup>9</sup>/L | 1.64 (1.16 – 2.30) | 0.005 |

Figure 1 illustrates the proportion of patients in the overall cohort who experienced recurrence or the composite outcome of recurrence or 30-day mortality, given the number of prognostic factors in the multivariable logistic regression model. At a score of 0 prognostic factors, the recurrence rate was 3.5% and the rate of recurrence or 30-day mortality was 10.5%. At a score of 1, 2, or 3 prognostic factors, the recurrence rate increased to 14.8%, 16.5%, and 17.9%, respectively. Similarly, at 1, 2, or 3 prognostic factors, the rate of the composite outcome of recurrence or 30-day mortality increased to 21.8%, 31.0%, and 48.7%, respectively.

**Figure 1.**
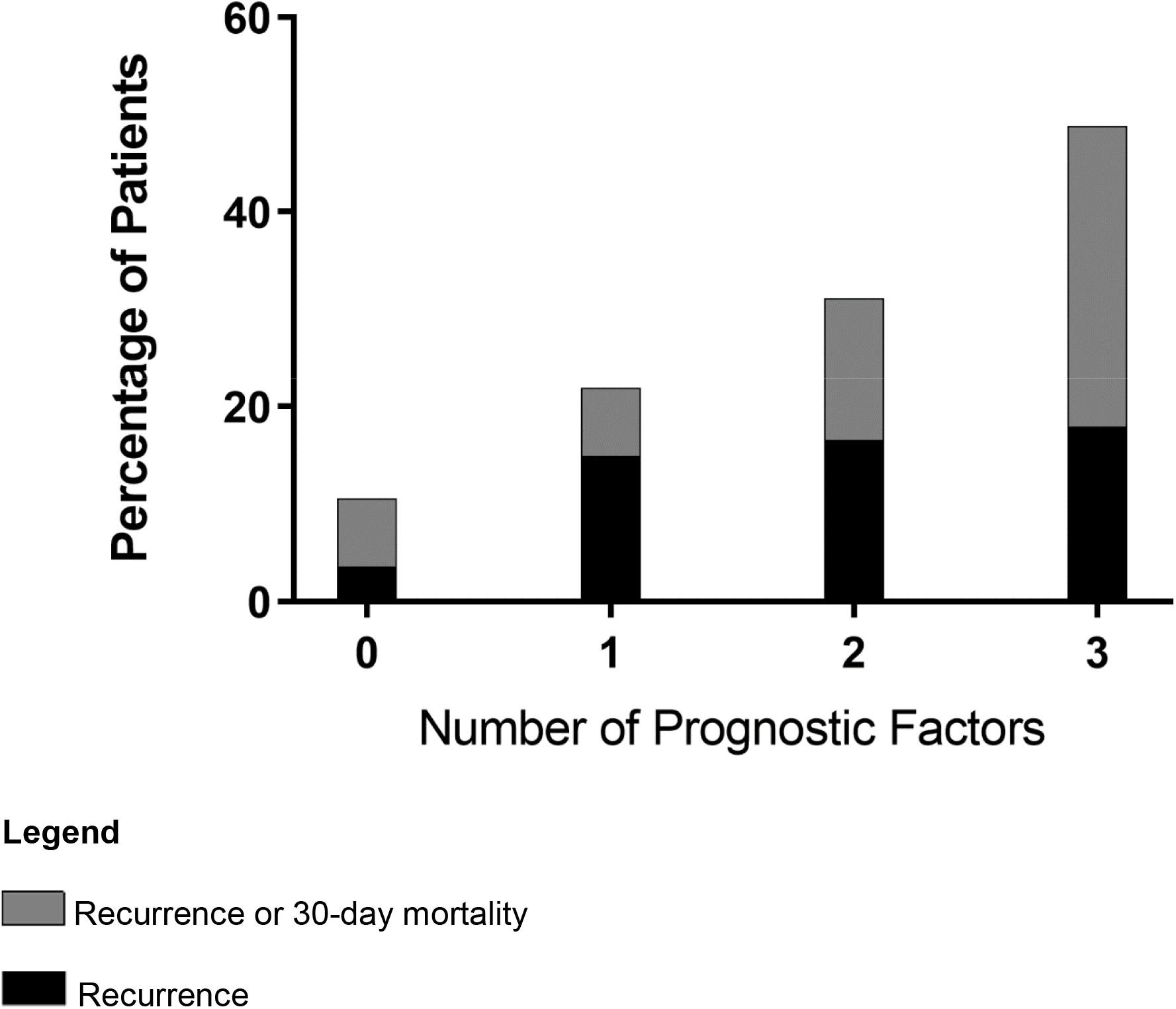
Proportion of patients in the overall cohort who experienced recurrence or the composite outcome of recurrence or 30-day mortality given the number of prognostic factors.

### Non-severe CDI cohort

There were 326 (44.2%) patients who were classified as having non-severe CDI, of whom 247 (75.8%) and 70 (21.5%) were treated with initial metronidazole and initial vancomycin, respectively. Among patients with first-episode non-severe CDI, recurrence, new incident infection, and 30-day mortality rates were 16.2%, 10.9%, and 5.3%, respectively, when treated with initial metronidazole, compared to 20.0%, 1.4%, and 10.0%, respectively, when treated with initial vancomycin. The use of initial vancomycin for treatment of first-episode non-severe CDI was associated with a reduction in new incident infection (OR_adj_: 0.11; 95% CI: 0.02–0.86; P=0.035) (Table 3).

**Table 3.** Predictors of new incident infection for patients with non-severe *Clostridioides difficile* infection.

| Variable | Univariable |  | Multivariable |  |
| --- | --- | --- | --- | --- |
|  | Odds Ratio (95% CI) | P | Odds Ratio (95% CI) | P |
| Initial vancomycin | 0.12 (0.02 – 0.89) | 0.038 | 0.11 (0.02 – 0.86) | 0.035 |
| Age ≥ 65 years |  |  | 0.56 (0.25 – 1.27) | 0.165 |
| Hospital-acquired infection |  |  | 1.36 (0.61 – 3.05) | 0.452 |

## Discussion

Although clinical trials have shown vancomycin to be superior to metronidazole for the treatment of severe CDI,^10^ some studies have suggested that metronidazole and vancomycin are equally effective for the treatment of non-severe CDI.^9–12^ Coupled with the higher cost of vancomycin and concerns associated with vancomycin-resistant enterococci, metronidazole has been the drug of choice for the treatment of non-severe CDI.^15^ Given that some guidelines recommend the use of metronidazole for non-severe CDI,^6–8^ Figure 1 illustrates a scoring rule which could help identify low-risk patient populations who are likely to do well on initial metronidazole.

In recent years, however, guidelines, such as those published by the IDSA and SHEA, have recommended initial fidaxomicin (preferred) or vancomycin (alternative) over metronidazole for the treatment of non-severe CDI.^5^ Our findings support the recommendation to use vancomycin when fidaxomicin is unavailable, which is applicable to institutions that rely on metronidazole and vancomycin for the treatment of CDI. A strength of our investigation was the long duration of follow-up, 26 weeks in a pragmatic real-world setting, which allowed for conclusions about the impact of initial vancomycin treatment on new incident infection, defined as a CDI episode occurring more than 8 weeks after the initial infection. In an adjusted analysis, the use of initial vancomycin for the treatment of first-episode non-severe CDI was associated with a reduction in new incident infection. At our institution, there has been an overall decline in new incident infection rates in recent years, though it is unclear whether this should be attributed to the increased use of initial vancomycin treatment or other factors. A possible explanation for the lower new incident infection rate experienced by patients who received initial vancomycin could be the broader spectrum antimicrobial activity of metronidazole which may be more disruptive to gut flora and potentially predispose patients for new incident infection.^2,16,17^

### Limitations

There were some limitations to our observational study. Patients were not randomized to treatment groups, for instance, and could have differed on key prognostic factors that may have influenced patient outcomes. To mitigate differences between groups, an adjusted analysis was done using prognostic factors found for the overall cohort. However, a limitation of our analysis was the exclusion of comorbidities in the database. Another limitation was the possible misclassification of exposures, since the treatment of CDI can evolve over a patient’s stay. Patients were analyzed in a manner analogous to the intention-to-treat approach, based on the initial treatment assignment, and any treatment switches after 72 hours were disregarded. Furthermore, although there was no observed increase in instances of vancomycin-resistant enterococci throughout our investigation, testing for this organism was non-systematic in nature. Finally, our investigation could have included some patients colonized with *Clostridioides difficile*, instead of true infection. This is unlikely, however, as diagnosis of CDI was confirmed by one of three experienced infectious disease physicians, all cases were followed by a dedicated CDI consultation team, and CDI diagnosis required the presence of a positive toxin assay together with a clinical presentation of 3 or more type 5–7 loose stools on the Bristol stool scale.

## Conclusions

Initial vancomycin was associated with a reduction in new incident infection in the treatment of adult inpatients with first-episode non-severe CDI. These findings support the use of initial vancomycin for all inpatients with CDI, when fidaxomicin is unavailable.

## Data Availability

The datasets generated for the study are not publicly available due to the presence of personally identifiable information. Aggregate data, however, are presented in the manuscript within the Tables and the Figure and are available from the corresponding author on reasonable request.

## Authors’ contributions

Kevin Zhang and Marek Smieja contributed to the conception and design of the work. All of the authors contributed to the acquisition of data, analysis, and interpretation of results. Kevin Zhang and Marek Smieja drafted the manuscript. All of the authors critically revised and approved the final manuscript.

## Ethics approval and consent to participate

This study was conducted in accordance with the Helsinki Declaration. The study protocol was approved by the Hamilton Integrated Research Ethics Board (Project: 2018-3543). The Hamilton Integrated Research Ethics Board has categorized this retrospective cohort study as minimal risk, defined as no potential for negative impact on the health and safety of the participant, and waiver of consent for participation was obtained.

